# Linguistic Dynamics: Women vs. the General Population in Reddit’s ADHD Discussions

**DOI:** 10.1101/2024.09.20.24314091

**Authors:** Muhammad Mahbubur Rahman

## Abstract

Understanding how language reflects the experiences of individuals with attention-deficit/hyperactivity disorder (ADHD) is important for developing targeted support strategies. This research investigates the linguistic patterns exhibited in discussions within women-centric and general ADHD population on Reddit, exploring how gender influences language use. By analyzing language, the study uncovers unique linguistic patterns and illuminates how women with ADHD express themselves, share experiences, and seek support compared to the general ADHD population. By leveraging deep learning-based embedding, clustering and named entity recognition, the study conducts a comprehensive analysis. Results reveal that women with ADHD emphasize themes like “ADHD Awareness and Diagnosis” and “Emotional Well-being”. Conversely, the general population highlights “Medication and Treatment”, and “Work and Academic Challenges”. Statistical tests confirm significant differences in both linguistics and emotional expression between the two groups, emphasizing the importance of gender considerations in understanding ADHD language nuances and tailoring interventions accordingly.

## 1. Introduction

Attention-deficit/hyperactivity disorder (ADHD) is a widespread neurodevelopmental condition that often begins in childhood and can potentially persist into adulthood, impacting approximately 9.8% of U.S. children[1] and 4.4% of adults[2]. Treating and managing adult ADHD presents distinct challenges compared to childhood cases, primarily due to several factors: overlapping symptoms with other adult mental illnesses, complex medical histories, difficulty accessing information on symptoms readily observable in children by parents and teachers, and the presence of misperceptions and stigma surrounding the disorder[3–6]. However, with the rise of social media platforms like Twitter (X) and Reddit, adults frequently share their personal experiences and emotion on these platforms which provide a valuable source of publicly available data for deeper understanding of adult ADHD. Some research has utilized social media data from platforms like Reddit and Twitter to understand and identify ADHD and other mental health conditions[7–10]. However, much of this research has focused primarily on disease detection, general language analysis, and comparative evaluations of methodologies, and often relies on self-reported data.

A critical gap exists in comprehensively analyzing language use on social media, particularly Reddit, specifically regarding discussions about ADHD between women and the general population. This study centers on Reddit data to explore the language discussed by women with ADHD and the general population, emphasizing linguistic patterns, emotional aspects, and conversations related to medication, therapy, and disorder itself. Data from two subreddits, WomenADHD and the general ADHD population, are utilized to uncover linguistic differences between women and the general population regarding ADHD-related discussions. Specific research questions and hypotheses are defined for this investigation, employing simple statistical hypothesis tests to gain insights into these aspects. The following are our research questions and hypotheses:

### Research Question 1 (RQ1)

What are the common linguistic patterns observed in language used by women with ADHD and the general ADHD population?

### Hypothesis 1

We hypothesize that linguistic differences will exist in language use between women with ADHD and the general ADHD population, with respect to distinct language patterns used by each group.

### Research Question 2 (RQ2)

Do women with ADHD express their emotional expressions in their language use that differ statistically from the general ADHD population?

### Hypothesis 2

We hypothesize that women with ADHD and individuals in the general ADHD population will display statistically differences in the way they express emotions within their language use.

### Research Question 3 (RQ3)

What medication, therapy, and disease entities are identified within the language used by women with ADHD compared to the general ADHD population, and what is the prevalence of these entities in each group’s discussions?

Our primary contributions are as follows:

- Examining gender-specific language patterns (i.e., between women and general population) in ADHD discussion sourced from social media platforms like Reddit.
- Analyzing emotional expressions, medication, therapy and disease among women and the general population.
- Uncovering both shared linguistic traits and women-specific nuances in the language used within ADHD communities.

Through an analysis of language use patterns, this study uncovers how women with ADHD express themselves and seek support within online communities. These insights can be valuable in developing gender-specific support strategies. Furthermore, this research seeks to enhance our understanding of the linguistic dynamics that characterize both women-centric and general ADHD communities. This understanding can shed light on potential factors influencing community engagement and cohesion.

**Figure 1** presents a visual representation of the overall study design.

**Fig. 1.**
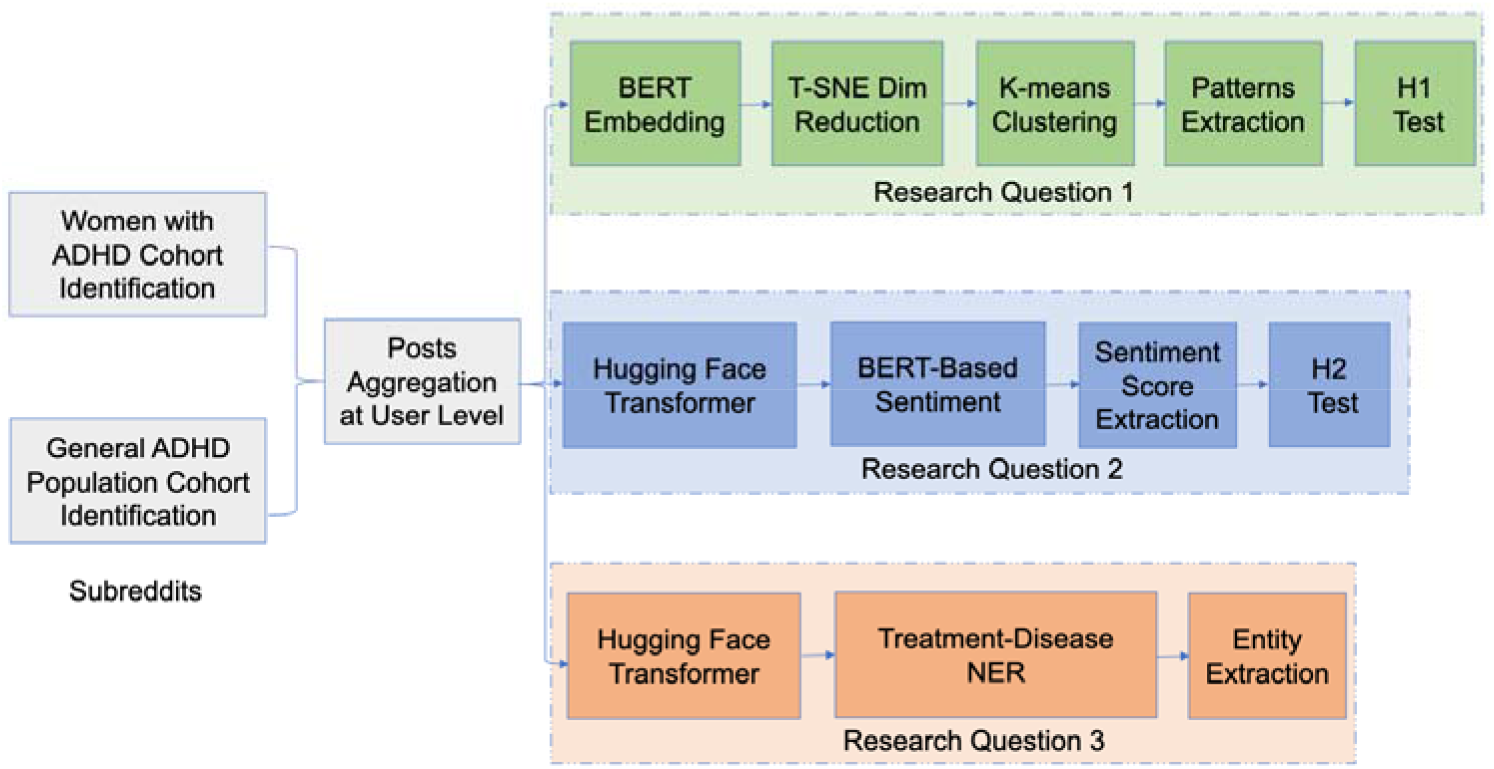
The overview of the whole study

## 2. Materials and Methods

### 2.1. Study data sample

To ensure representativeness, a random sample of 10,000 unique users was drawn from the ADHD Women subreddit, representing the population of women with ADHD. A separate random sample of 10,000 unique users was obtained from the general ADHD subreddit. To prevent potential overlap between the two groups, user accounts found in both subreddits were excluded prior to sampling. Subsequently, all posts authored by the selected users in both groups were chosen. For privacy considerations, the original usernames associated with each post were anonymized using a SHA-256 hash function. This process irreversibly replaces usernames with unique hash values, ensuring the anonymity of the original authors. A separate mapping table was created to maintain a link between the original usernames and their corresponding hash values for potential future reference. Finally, the collected posts were concatenated at the user level. This step aims to capture user-specific linguistic patterns rather than focusing solely on individual posts.

### 2.2. Data analysis

To address ***RQ1***, which focuses on identifying linguistic patterns, we used a deep learning-based word embedding approach known as Bidirectional Encoder Representations from Transformers (BERT)[11]. This pre-trained BERT model generated a numerical representation (embedding) for each user’s Reddit posts within our 20,000-user cohort encompassing both women with ADHD and the general ADHD population. Each user’s posts sequence was transformed into a 512-dimensional vector embedding. Prior to embedding, necessary preprocessing and data cleaning steps, including the removal of stop words, punctuation, and lemmatization were applied. Following the embedding representation, a dimensionality reduction technique using t-distributed Stochastic Neighbor Embedding (T-SNE)[12] was applied to the 512-dimensional embedding matrix for each user’s posts, resulting in a 2-dimensional matrix. T-SNE was applied with 5000 iterations in a multi-core setup, facilitating data visualization and clustering. Subsequently, K-means clustering was implemented to cluster the embeddings of women with ADHD and the general population with ADHD separately. The clustering models were trained with 5000 iterations, aiming to identify 50 distinct clusters within each group.

For each cluster within each population group, representative original Reddit posts were retrieved, and term frequency-inverse document frequency (TF-IDF[13]) techniques were applied to extract n-gram linguistic patterns. The TF-IDF vectorizer was trained using unigram, bigram, and trigram language models. Terms with TF-IDF scores were extracted and presented visually using a word cloud to interpret linguistic patterns between the two groups.

To investigate ***RQ2***, which centers on emotional expression within the data, we utilized sentiment analysis techniques based on the BERT language model. This approach enabled us to automatically assess the sentiment polarity of user-level aggregated Reddit posts for both women with ADHD and the general ADHD population. Sentiment scores were assigned on a scale of 1 to 5 stars, where 1 represented extremely negative sentiment and 5 represented extremely positive sentiment. Scores of 2 and 4 indicated slightly negative and slightly positive sentiment, respectively, while a score of 3 denoted neutral sentiment.

By utilizing sentiment analysis, we aimed to gain insights into how individuals with ADHD discuss their experiences on the Reddit platform and express emotions related to ADHD through their language use. Leveraging the Hugging Face Transformers-based pipeline, we utilized a publicly available pretrained language model, ‘*nlptown/bert-base-multilingual-uncased-sentiment*[14], which was fine-tuned for sentiment analysis tasks. The chosen model utilizes a BERT architecture with key characteristics including 512-dimensional embeddings, 12 attention heads, 12 hidden layers, a dropout rate of 0.1, and a vocabulary size of 105,879. These characteristics contribute to the model’s ability to effectively capture semantic relationships within the text and assign appropriate sentiment scores. By applying this pre-trained model, we computed sentiment scores for the aggregated posts at the user level within both population groups.

To address ***RQ3***, we explored the language of women with ADHD and the general ADHD population, focusing on extracting entities related to medication, therapy, and diseases from aggregated posts at the user level. Leveraging a pre-trained treatment-disease Named Entity Recognition (NER) model accessible on the Hugging Face platform[15], specifically designed for detecting treatment and disease entities in health-related text, we conducted our analysis. The chosen model is a token classification model with key characteristics including 12 hidden layers, a hidden layer dropout rate of 0.1, a 512-dimensional embedding size, 12 attention heads, and a vocabulary size of 30,522. These characteristics enable the model to effectively analyze individual tokens within the text and assign appropriate entity labels. The model utilizes five class labels:

B-d (beginning of a disease mention), I-d (intermediate mention of a disease), B-t (beginning of a treatment mention), I-t (intermediate mention of a treatment), and O (any other mention). Our extraction process involved identifying relevant mentions, and for entities with intermediate mentions, we concatenated them with their corresponding beginning mention to form complete entities. For instance, “Cognitive Behavioral Therapy” is a complete entity, with “Cognitive” as the beginning mention and “Behavioral” and “Therapy” as intermediate mentions. After successfully extracting all treatment and disease-related entities, we calculated the prevalence of these entities within each population group to gain insights into the focus of health-related discussions.

### 2.3. Statistical analysis

To assess ***hypothesis 1***, an independent t-test was conducted between the two sets of clusters derived from the distinct ADHD population groups. As the clusters were computed independently from two sets of Reddit posts from separate ADHD populations, they were treated as independent entities. The centers of each cluster within each ADHD population group were extracted, and T-SNE dimensionality reduction was applied once again to obtain a singular central point for each cluster. Subsequently, the statistic t-test was used to test the ***hypothesis 1*** between the centers of the two sets of clusters for the two ADHD population groups. To test ***hypothesis 2***, we conducted a statistical t-test between sentiment scores of two groups of ADHD population.

We used Python 3.9 for all analyses. Pandas and TensorFlow libraries[16] were utilized for data processing and embedding representation. The scikit-learn[17] machine learning library facilitated clustering and TF-IDF vectorization, while the T-SNE library handled dimensionality reduction. Results visualization was conducted using the Python WordCloud library and Matplotlib. Sentiment analysis and Named Entity Recognition (NER) detection were performed using Python transformers from Hugging Face. The statistical t-test was conducted using the SciPy[18] stats library.

### 2.4. Ethics Statement

This research used publicly available data from ADHD discussions on Reddit. There were no interactions with users involved in the study. Given this, the research was classified as exempt from Institutional Review Board approval due to the absence of human participants. However, there may still ethical concerns with using these data. While users within ADHD-related subreddits implicitly share their experiences on the platform, such sharing does not constitute explicit consent for research analysis. Additionally, while personally identifiable information (PII) is not shared by the users on these discussions, established ethical guidelines[19] are followed to ensure anonymity. Therefore, direct usernames and any other potentially identifying information are not reported within this study. This approach serves to protect the privacy of individuals whose posts were included in the study.

## 3. Results

### 3.1. Linguistic patterns

An exploration of the ten most frequent terms used by the women with ADHD cohort (*meds, diagnosis, help, ADHD, feel like, advice, work, feeling, ADHD meds, and tips*) reveals key areas of focus. These terms center on medication experiences, seeking support and guidance related to ADHD, emotional well-being, and navigating challenges in work and daily life. For example, words or phrases like *“meds”, “diagnosis” and “ADHD meds”* may suggest a focus on medication use and potentially managing treatment. Additionally, terms like *“help” and “advice”* may indicate a desire for support and guidance, potentially from peers. Further investigation of the 50 identified clusters within the women with ADHD group reinforces these areas of focus. The clusters highlight themes surrounding various aspects of ADHD such as **ADHD Awareness and Diagnosis** (terms: *ADHD, diagnosed, finally got diagnosis, newly diagnosed*), **Medication and Treatment** (terms: *meds, medication, adhd meds, prescribed adderall, vyvanse, adderall xr*), **Seeking Support and Advice** (terms: *help, need help, need advice, looking for advice*), **Emotional Well-being** (terms: *feel, like, feeling like, really struggling, imposter syndrome*), and **Work and Life Challenges** (terms: *work, life, struggling work, dealing, hard time*). Detailed TF-IDF scores for each cluster in the women with ADHD population are available in the **supplemental material** (*women_top_tfidf_scores_per_cluster*.*xlsx*). **Figure 2** presents a word cloud based on the TF-IDF scores, showcasing an overview of all clusters for women with ADHD. Individual word clouds for each cluster in the women with ADHD group are given in the **supplemental material** (*wordcloud_women*.*docx*).

**Fig. 2.**
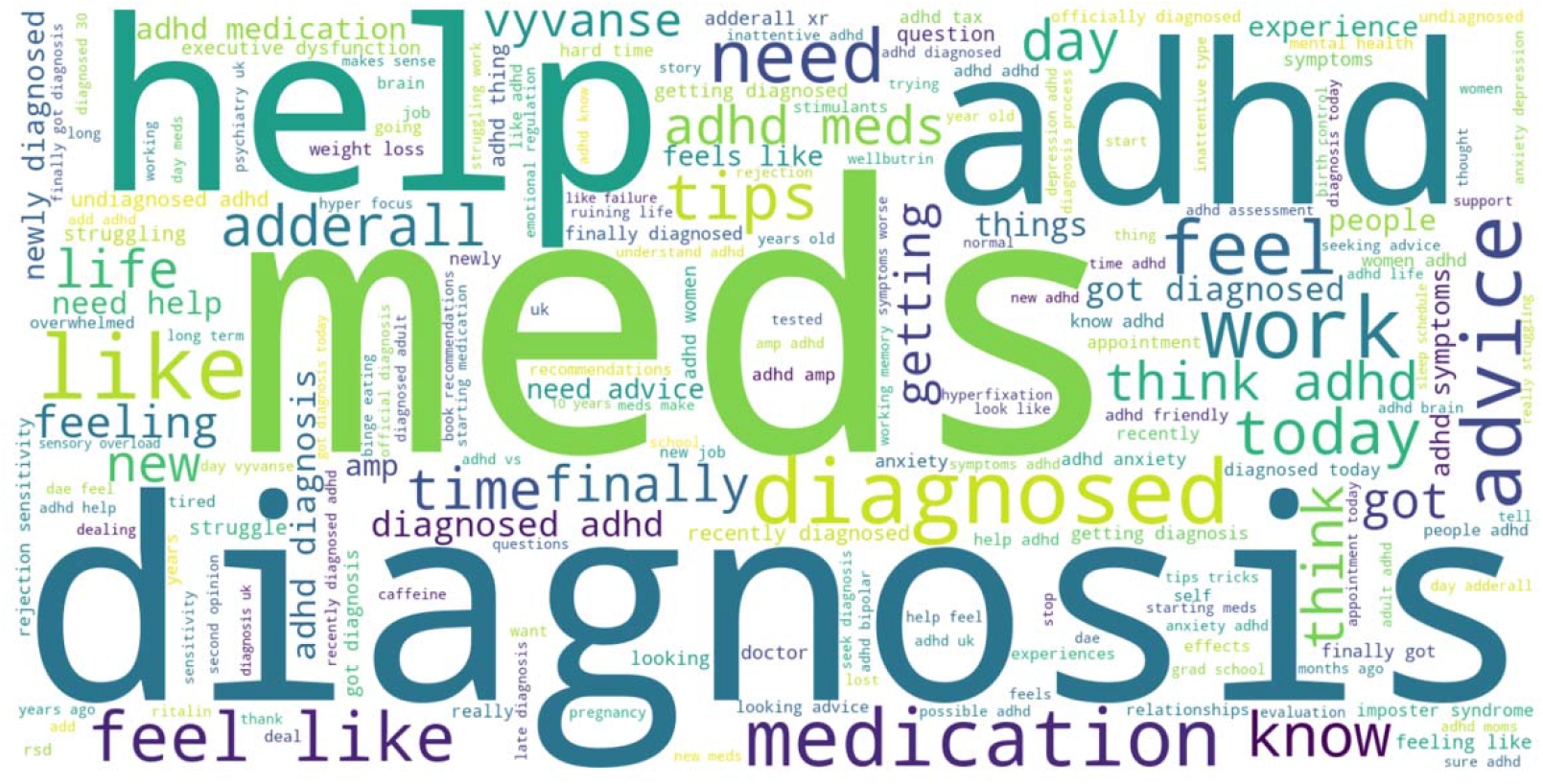
Word cloud for women with ADHD

An examination of the ten most frequent terms used by the general population with ADHD (*meds, Adderall, medication, help, Vyvanse, advice, diagnosed, people, Ritalin and newly diagnosed*) reveals key areas of focus. These terms center on medication use, with specific mentions of medications like *“Adderall”, “Vyvanse” and “Ritalin”* alongside a desire for help and support *(“help”, “advice”*). Additionally, terms like *“diagnosed” and “newly diagnosed”* suggest experiences with the diagnostic process. Further analysis of the 50 identified clusters within the general ADHD group reinforces these areas of focus, while also highlighting other aspects of living with ADHD. The clusters capture distinct themes emerged, including **Medication and Treatment** (terms: *medication, meds, vyvanse, adderall, strattera, ritalin*), **Work and Academic Challenges** (terms: *job, new job, career choices, finished homework*), **Coping Mechanisms** (terms: *coping mechanisms, self-help, life improvement*), **Impact on Relationships** (terms: *friends, family, relationships, best friend*), and **Social Interaction** (terms: *telling people, hard time communicating, social challenges*). Detailed TF-IDF scores for each cluster in the general ADHD population are provided in the **supplemental material** (*general_top_tfidf_scores_per_cluster*.*xlsx*). **Figure 3** presents a consolidated word cloud for all 50 clusters in the general population. Individual word clouds for each cluster in the general ADHD group can be accessed in the **supplemental material** (*wordcloud_general*.*docx*).

**Fig. 3.**
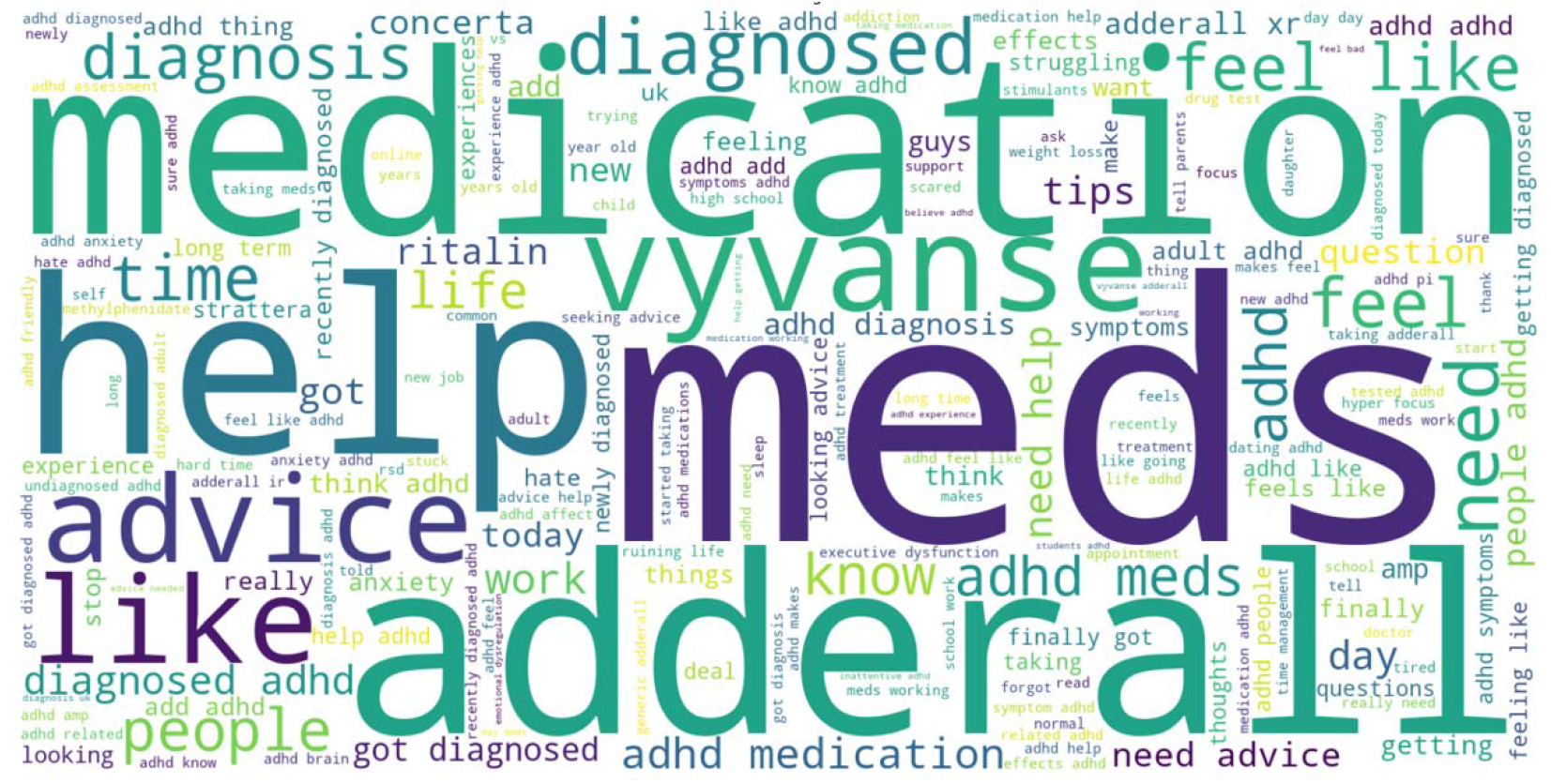
Word cloud for general ADHD population

To investigate potential linguistic differences in language use between women with ADHD and the general ADHD population (**hypothesis 1**), we conducted an independent statistical t-test. This analysis compared the center points (centroids) of the identified clusters for each group. The results revealed a statistically significant difference (t-statistic = 10.63, p-value < 0.000051), suggesting distinct linguistic characteristics in the discussions within these populations. **Figure 4** presents bell curve visualizations of cluster centers for each ADHD population group, demonstrating normal distributions in both cases.

**Fig. 4.**
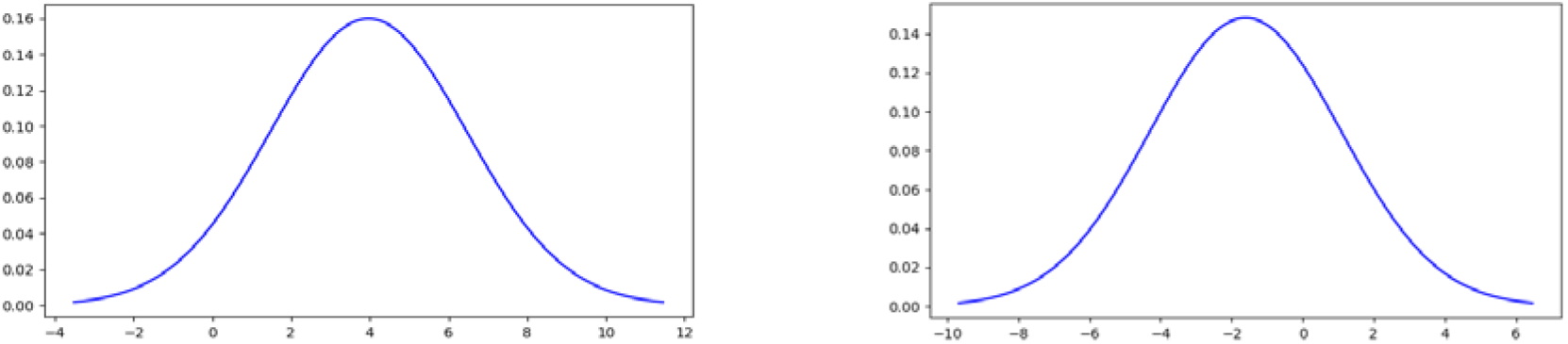
Bell curves of cluster centers for women (left) and general (right) ADHD groups

### 3.2. Emotional expression

To assess emotional expression within the language used by participants, sentiment scores, ranging from 1(negative) to 5(positive), were calculated for each individual in both the women with ADHD and general ADHD population groups using the pretrained language model *bert-base-multilingual-uncased-sentiment*. Detailed sentiment scores for all users in each group are provided in the **supplemental material** (*women_sentiment*.*xlsx* and *general_sentiment*.*xlsx*). To ensure user privacy, author names were anonymized using SHA-256 hashing in the provided sentiment scores in the supplemental material.

A statistical t-test was conducted to investigate potential differences in emotional expression between the two groups in their language use (**hypothesis 2**). The analysis revealed a statistically significant difference (t-statistic = 6.49, p-value <0.000089) in the sentiment scores, suggesting distinct emotional patterns within the language used by women with ADHD compared to the general ADHD population. The substantial effect size, indicated by the t-statistic, strengthens this finding.

### 3.3. Medication, therapy, and diseases

In the women with ADHD group, prominent medication entities include *stimulants, antidepressants, hydroxyzine, methylphenidate, atomoxetine, Wellbutrin, and sertraline*. Top disease entities within this group encompass *anxiety, depression, autism, insomnia, migraine, and addiction*. Similarly, within the general ADHD population, notable medication entities include *methylphenidate, Ritalin, Adderall, music, stimulant medication, antidepressants, and marijuana*. Prevailing disease entities for this population include *anxiety, depression, insomnia, autism, addiction, and psychosis*. A summarized list of 20 entities for each group is provided in **Table 1**, while a detailed list of medication, therapy, and disease entities with their prevalence is available in the **supplemental material** (*women_medication*.*docx, women_disease*.*docx, general_medication*.*docx*, and *general_disease*.*docx*).

**Table 1:**
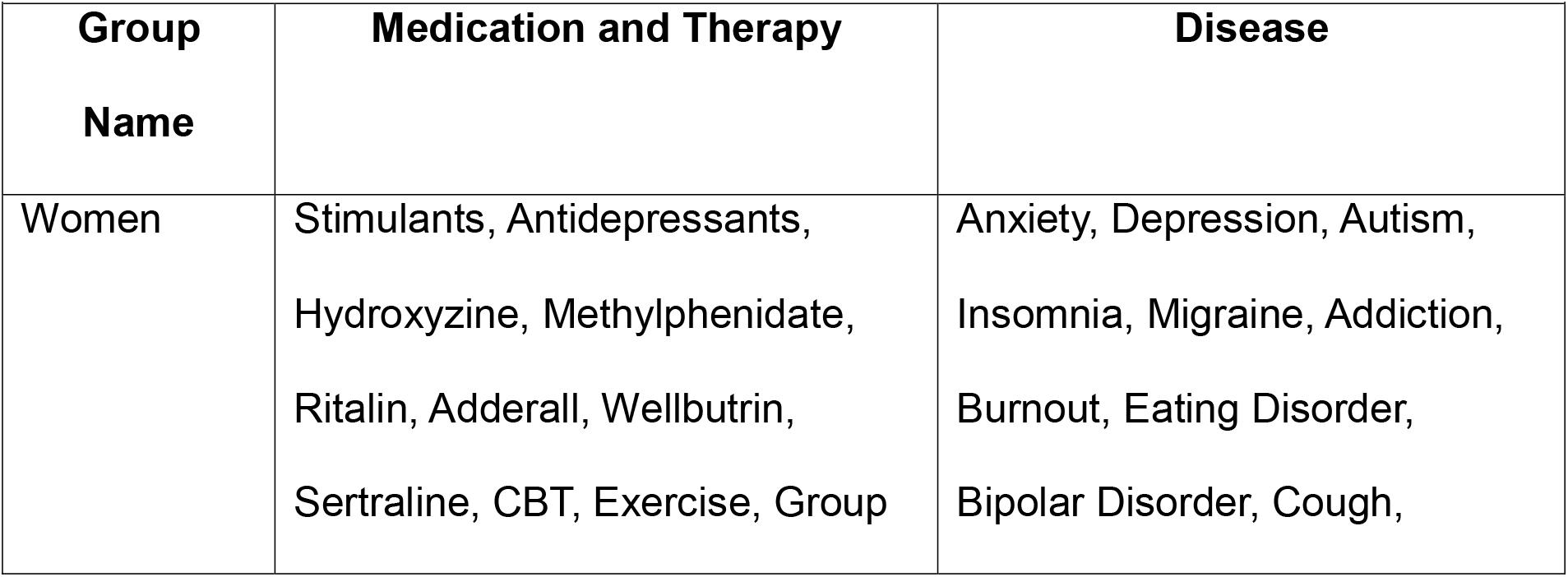
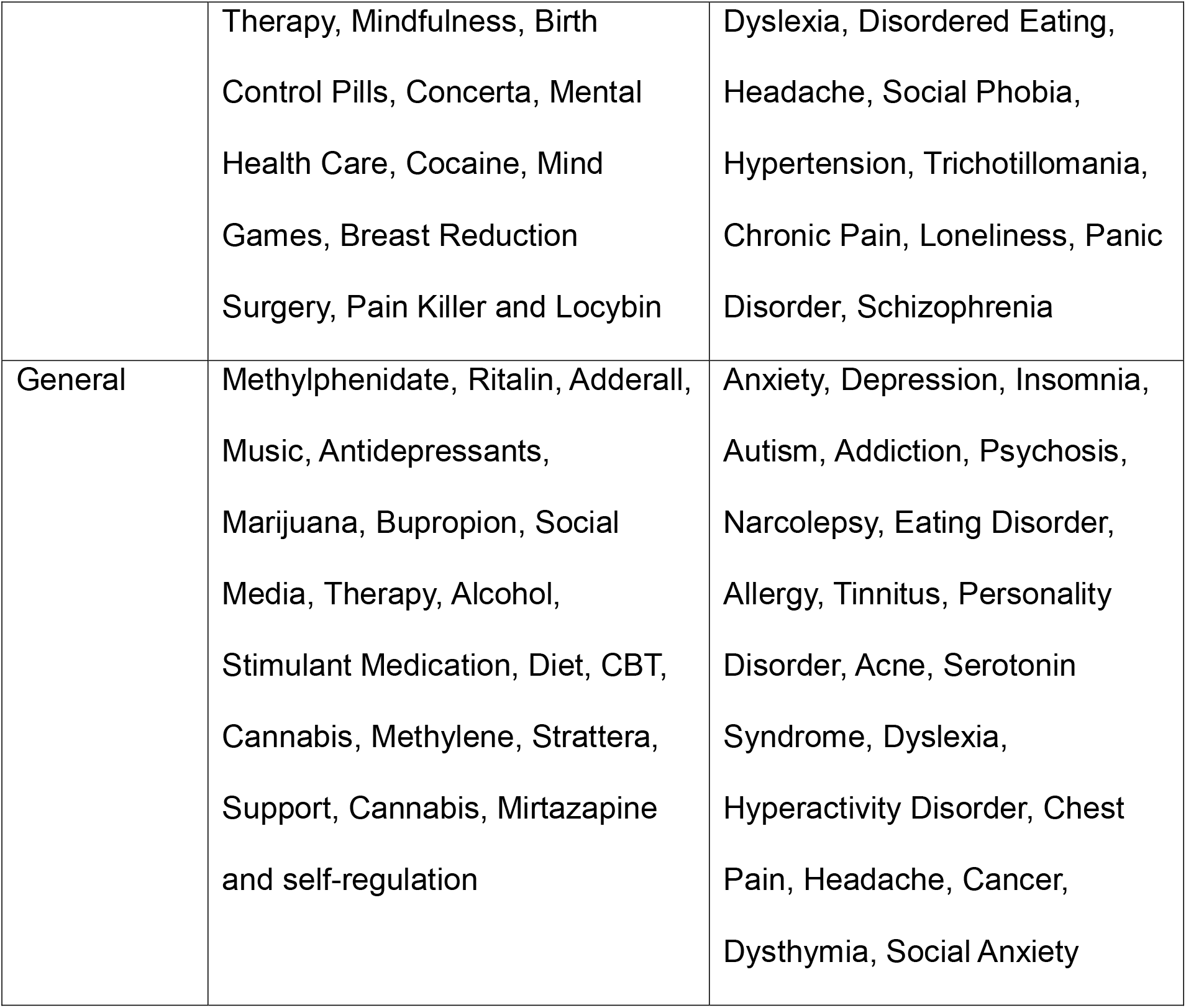
A list 20 of entities

| Table 1: A list 20 of entities |  |  |
| --- | --- | --- |
| Group Name | Medication and Therapy | Disease |
| Women | Stimulants, Antidepressants, Hydroxyzine, Methylphenidate, Ritalin, Adderall, Wellbutrin, Sertraline, CBT, Exercise, Group | Anxiety, Depression, Autism, Insomnia, Migraine, Addiction, Burnout, Eating Disorder, Bipolar Disorder, Cough, |
|  | Therapy, Mindfulness, Birth Control Pills, Concerta, Mental Health Care, Cocaine, Mind Games, Breast Reduction Surgery, Pain Killer and Locybin | Dyslexia, Disordered Eating, Headache, Social Phobia, Hypertension, Trichotillomania, Chronic Pain, Loneliness, Panic Disorder, Schizophrenia |
| General | Methylphenidate, Ritalin, Adderall, Music, Antidepressants, Marijuana, Bupropion, Social Media, Therapy, Alcohol, Stimulant Medication, Diet, CBT, Cannabis, Methylene, Strattera, Support, Cannabis, Mirtazapine and self-regulation | Anxiety, Depression, Insomnia, Autism, Addiction, Psychosis, Narcolepsy, Eating Disorder, Allergy, Tinnitus, Personality Disorder, Acne, Serotonin Syndrome, Dyslexia, Hyperactivity Disorder, Chest Pain, Headache, Cancer, Dysthymia, Social Anxiety |

## 4. Discussion

While the analysis revealed distinct clusters within the women with ADHD cohort and the general ADHD population, some commonalities emerge in their discussions. Both groups share concerns about medication with terms like *“medication”, “vyvanse” and “adderall”* indicating a shared interest in pharmacological approaches to managing ADHD symptoms. Seeking help and support is a prevalent theme across both groups, as reflected in terms like *“help”, “need help” and “advice”*. Personal experiences and reflections are key linguistic patterns in both clusters, with terms such as *“feel”, “like” and “feel like”* suggesting shared emotional expressions.

The most significant distinctions between women with ADHD and the general ADHD population lie in the themes and linguistic patterns within their identified clusters. In clusters specific to women with ADHD, themes such as “*ADHD Awareness and Diagnosis*”, “*Medication and Treatment*”, “*Seeking Support and Advice*”, “*Emotional Well-being*” and “*Work and Life Challenges*” take precedence. Women with ADHD express a strong focus on awareness, emotional struggles, and the unique impact of ADHD on their professional and personal lives. The linguistic terms associated with these themes, such as *“diagnosed”, “meds”, “help”, and “feel”* further reflect the distinct experiences and challenges faced by women with ADHD.

On the other hand, clusters representing the general population with ADHD highlight themes like “*Medication and Treatment*”, “*Work and Academic Challenges*”, “*Coping Mechanisms*”, “*Impact on Relationships*” and “*Social Interaction*”. The general population places greater emphasis on pharmaceutical solutions, vocational difficulties, coping strategies, the influence of ADHD on relationships, and challenges in social interactions. Linguistic terms such as *“medication”, “job”, “coping mechanisms”, “friends” and “telling people”* highlight these differences, indicating a broader range of concerns and perspectives within the general ADHD population.

These differences suggest that while there are commonalities in the challenges associated with ADHD, the nuanced experiences and priorities differ between women with ADHD and the general population.

The statistical analysis provides compelling support for **hypothesis 1**. A t-statistic of 10.63 and a highly significant p-value less than 0.000051 offer strong evidence for distinct linguistic patterns in the language used between women with ADHD and the general ADHD population. This suggests that women with ADHD usually use a unique set of language features and terminology compared to the general ADHD group.

Our investigation whether women with ADHD express emotional expressions differently compared to the general ADHD population provide insights into the nuanced dynamics of language use within these groups. The observed statistically significant difference identified through the t-test, with a substantial t-statistic of 6.49, shows the presence of meaningful distinctions in the emotional content of language. This finding strongly supports our **hypothesis 2**, suggesting that there are unique patterns in how individuals with ADHD, especially women, articulate their emotions compared to the general ADHD population. The significant p-value less than 0.000089 also supports the hypothesis, emphasizing the importance of considering gender-specific language expressions within the ADHD community. These findings provide a signal of how language serves as a channel for emotional expression among individual with ADHD from different genders. Recognizing these distinctions could inform tailored interventions, support mechanisms, and educational strategies that account for the unique communication needs within the ADHD community.

In examining the medication landscape, both women with ADHD and the general population with ADHD share common ground in their reliance on traditional pharmacological interventions. Stimulants like *methylphenidate, Ritalin, and Adderall* are prevalent in discussions from both groups, indicating a collective dependence on conventional medical treatments for managing ADHD symptoms. This shared trend highlights a universal inclination towards established medications within the ADHD community. Furthermore, both populations face prevalent mental health concerns, with *anxiety and depression* emerging as predominant challenges. This parallel mental health burden suggests a shared psychological impact of ADHD, irrespective of gender. The discussions around these mental health entities serve as a common thread, emphasizing the need for holistic mental health considerations in both groups.

Despite these similarities, important dissimilarities exist, particularly in the realm of therapeutic approaches and disease focus. Women with ADHD exhibit a unique inclination towards *group therapy*, indicating a potential preference for collaborative therapeutic settings. In contrast, the general ADHD population displays a broader range of non-traditional approaches, including discussions around *music, marijuana, and diet*. This diversity implies a willingness to explore unconventional methods beyond standard medical practices, possibly influenced by the heterogeneous nature of the general ADHD population. Moreover, the prevalence of *autism-related discussions* is significantly higher among women with ADHD, highlighting a gender-specific concern for neurodevelopmental issues. This distinct focus emphasizes the need for gender-specific considerations in understanding and addressing comorbidities associated with ADHD.

While shared trends provide a foundation for general ADHD medication and diseases, recognizing and understanding these gender-specific nuances is crucial for tailoring interventions effectively. These dissimilarities not only reflect individual preferences but also emphasize the importance of acknowledging diverse needs within the ADHD community. Further research is needed to investigate the reasons behind these variations and their implications for healthcare delivery and outcomes.

## 5. Limitation

Using Reddit data may introduce potential bias, especially given the absence of demographic information such as age, location, and race. Ideally, a more comprehensive analysis could be conducted with access to such data. Additionally, the lack of a dedicated subreddit specifically for men with ADHD necessitated limiting our analysis to a comparison between women with ADHD and the general ADHD population. Moreover, the NER models used for extracting medication, therapy and disease entities may incur misclassifications, which should be considered when interpreting our findings.

## 6. Conclusion

This study investigated the language patterns used in discussions about ADHD on Reddit, focusing on a comparison between women with ADHD and the general ADHD population. While both groups shared concerns, such as medication use and seeking support, statistically significant linguistic differences emerged. Discussions by women with ADHD emphasized themes like “ADHD Awareness and Diagnosis” suggesting a deeper focus on understanding their condition. Conversely, the general ADHD population’s discussions prioritized themes like “Work and Academic Challenges”.

These findings highlight the need for tailored interventions that consider gender-specific linguistic variations in communication approaches. Additionally, the study provided insights into medication use and potential mental health concerns expressed within these online communities. This research contributes to a more nuanced understanding of how ADHD reveals differently for women compared to the general ADHD population. By emphasizing the importance of recognizing and addressing gender-specific language dynamics, this study opens doors for developing more targeted support strategies.

## Data Availability

All data produced are available online at Reddit Platform

https://www.reddit.com

