## Supplemental for "Linguistic Dynamics: Women vs. the General Population in Reddit’s ADHD Discussions": general_disease.docx

{

"anxiety": 135,

"depression": 112,

"insomnia": 21,

"autism": 13,

"addiction": 10,

"migraine": 7,

"bipolar disorder": 6,

"anxiety disorder": 6,

"psychosis": 5,

"narc ole psy": 5,

"eating disorders": 4,

"eating disorder": 4,

"allergy": 4,

"tinnitus": 4,

"personality disorder": 3,

"acne": 3,

"serotonin syndrome": 2,

"dyslex ia": 2,

"hyperactivity disorder": 2,

"chest pain": 2,

"headache": 2,

"cancer": 2,

"mental illness": 2,

"anxiety medication": 2,

"sleep": 2,

"back pain": 2,

"dysthymia": 2,

"mental disorder": 1,

"anxiety psychiatr": 1,

"adhd work": 1,

"social anxiety": 1,

"sera ton in syndrome": 1,

"hair loss": 1,

"decay": 1,

"tachycardia": 1,

"adhd medicine": 1,

"dizziness": 1,

"anorexia": 1,

"allergies": 1,

"heart failure": 1,

"burnout": 1,

"narcisism": 1,

"speech delay": 1,

"constipation": 1,

"bulimia": 1,

"headaches": 1,

"neck pain": 1,

"mania": 1,

"cardiovascular disease": 1,

"celiac disease": 1,

"asthma": 1,

"mood disorder": 1,

"fetal alcohol spectrum disorder": 1,

"urinary tract infections": 1,

"substance": 1,

"chronic pain": 1,

"anxiety support": 1,

"ptsd": 1,

"substance use disorder": 1,

"sleep paralysis": 1,

"insomnia medication": 1,

"schiz oid personality disorder": 1,

"iron deficiencies": 1,

"psychiatry": 1,

"sexual dysfunction": 1,

"sleepiness": 1,

"trichotillomania": 1,

"adhd diagnosis": 1,

"social pho bia": 1,

"tourette": 1,

"adhd drugs": 1,

"alcohol use disorder": 1,

"parkinson's disease": 1,

"blindness": 1,

"mental": 1,

"major depressive disorder": 1,

"dyslex": 1,

"sleep apnea": 1,

"substance abuse": 1,

"bipolar disorder er": 1,

"divertic uli tis": 1,

"depressive disorder": 1,

"hyperlipidemia": 1,

"vision disorder": 1,

"hypothyroidism": 1

}
