## Supplemental for "Linguistic Dynamics: Women vs. the General Population in Reddit’s ADHD Discussions": general_medication.docx

{

"medication": 103,

"methylphenidate": 24,

"stimulants": 20,

"ritalin": 15,

"adhd medication": 14,

"adderall": 14,

"music": 9,

"meds": 7,

"medications": 6,

"stimulant medication": 6,

"antidepressants": 4,

"marijuana": 4,

"atomoxetine": 4,

"wellbutrin": 3,

"bupropion": 3,

"social media": 3,

"therapy": 3,

"alcohol": 3,

"adoctor": 3,

"amphetamines": 3,

"adhd treatment": 3,

"caffeine": 3,

"diet": 3,

"treatment": 3,

"stimulant": 2,

"medicine": 2,

"cannabis": 2,

"methylphen": 2,

"a stimulant": 2,

"rit": 2,

"behavioral therapy": 2,

"dexedrine": 2,

"mirtazapine": 2,

"stress": 2,

"meditation": 2,

"strattera": 2,

"support": 2,

"a medication": 2,

"coffee": 2,

"concert a": 2,

"primary care": 2,

"self-regulation": 1,

"food": 1,

"ssri": 1,

"methylphenidat": 1,

"methylphenidatephen": 1,

"medication efficacy": 1,

"cognitive-behavioral therapy": 1,

"appropriate medication": 1,

"opiates": 1,

"doctors": 1,

"clonidine": 1,

"maths": 1,

"add medications": 1,

"metilphenidate": 1,

"multiple medications": 1,

"apsychiatrist": 1,

"video": 1,

"drug abuse": 1,

"your medication": 1,

"guanfa": 1,

"dsm-v": 1,

"self care": 1,

"a treatment plan": 1,

"focalinxr": 1,

"fluoxetine": 1,

"iv ketamine infusions": 1,

"vyvanse": 1,

"probiotic": 1,

"dopamine": 1,

"therapy abuse": 1,

"a screening test": 1,

"that video game": 1,

"lyrica": 1,

"pharmacological treatments": 1,

"trauma": 1,

"ethylphenendate": 1,

"methylphenendate": 1,

"anxiety treatment": 1,

"drug treatment": 1,

"toronto": 1,

"parenting": 1,

"group therapy": 1,

"the internet": 1,

"gene therapy": 1,

"another medication": 1,

"addiction": 1,

"psychotherapy": 1,

"university medication": 1,

"mental health treatment": 1,

"attention support": 1,

"resources": 1,

"mph": 1,

"stimulant drugs": 1,

"pain management": 1,

"melatonin": 1,

"dopamine levels": 1,

"sleep": 1,

"phone apps": 1,

"cbttherapy": 1,

"apps": 1,

"anapp": 1,

"no treatment": 1,

"citalopram": 1,

"setraline": 1,

"ginkgobiloba": 1,

"evaluated": 1,

"cbt": 1,

"aneuropsychologist": 1,

"alternative medicine treatments": 1,

"cbd": 1,

"my medication": 1,

"orange juice": 1,

"wearable": 1,

"self-medication": 1,

"tova": 1,

"atest": 1,

"concert": 1,

"drug test": 1,

"methylphenidatepressure": 1,

"non-med treatments": 1,

"regular": 1,

"motivational aid": 1,

"natural treatment": 1,

"tv": 1,

"assessment": 1,

"placebo": 1,

"dialectical behavior therapy": 1,

"neuropsych": 1,

"therapy sessions": 1,

"neuropsychological testing": 1,

"modafini": 1,

"anti-depressants": 1,

"breastfeeding": 1,

"ssi": 1,

"seeked treatment": 1,

"diagnosis": 1,

"mindfulness-based therapy": 1,

"metadate": 1,

"best treatment": 1,

"ice cream": 1,

"psychiatrist": 1,

"antidepressant": 1,

"no medication": 1,

"natural supplements": 1,

"cognitive behavioural therapy": 1,

"instruments": 1,

"vitamin c": 1,

"gad": 1,

"parenting advice": 1,

"online services": 1,

"the reward system": 1,

"app": 1,

"a psychiatric review": 1,

"video games": 1,

"stimulation": 1

}
