## Supplemental for "Linguistic Dynamics: Women vs. the General Population in Reddit’s ADHD Discussions": women_disease.docx

{

"anxiety": 137,

"depression": 98,

"autism": 22,

"insomnia": 13,

"migraine": 9,

"addiction": 9,

"burnout": 5,

"anxiety disorder": 4,

"eating disorder": 4,

"bipolar disorder": 4,

"cough": 3,

"dyslexia": 3,

"disordered eating": 3,

"headache": 3,

"eating disorders": 3,

"social phobia": 2,

"hypertension": 2,

"trichotillomania": 2,

"chronic pain": 2,

"anxiety medication": 2,

"loneliness": 2,

"panic disorder": 2,

"type 1 diabetes": 2,

"schizophrenia": 1,

"serotonin syndrome": 1,

"pelvic pain": 1,

"suicidal": 1,

"epilepsy": 1,

"dehydration": 1,

"dermatilliomania": 1,

"mental illness": 1,

"autism diagnosis": 1,

"endometriosis": 1,

"hyperhidrosis": 1,

"fibromyalgia": 1,

"sleep disorder": 1,

"eczema": 1,

"meniere's disease": 1,

"autism test": 1,

"attention deficit disorder": 1,

"constipation": 1,

"ptsd": 1,

"heart attack": 1,

"liver failure": 1,

"melasm": 1,

"generalized anxiety disorder": 1,

"body dysmorphic disorder": 1,

"tinnitus": 1,

"narcolepsy": 1,

"hyster": 1,

"pho": 1,

"adhd depressant": 1,

"bipolar depression": 1,

"panic": 1,

"celiac disease": 1,

"phobia": 1,

"hyperactivity disorder": 1,

"diabetes": 1,

"sleep paralysis": 1,

"erectile dysfunction": 1,

"dysmorphic disorder": 1,

"alcoholism": 1,

"menopause": 1,

"varicose veins": 1,

"alcohol abuse": 1,

"substance abuse": 1,

"blood pressure": 1,

"drug abuse": 1,

"iron deficiency": 1,

"psoriasis": 1,

"alcohol dependence": 1,

"dementia": 1,

}
