## Supplemental for "Linguistic Dynamics: Women vs. the General Population in Reddit’s ADHD Discussions": women_medication.docx

{

"medication": 65,

"stimulants": 9,

"therapy": 8,

"methylphenidate": 6,

"adhd medication": 6,

"treatment": 6,

"antidepressants": 5,

"ritalin": 4,

"atomoxetine": 3,

"stimulant medication": 3,

"meds": 3,

"adhd treatment": 3,

"adderall": 3,

"wellbutrin": 3,

"social media": 2,

"diet": 2,

"sertraline": 2,

"rit": 2,

"music": 2,

"bupropion": 2,

"caffeine": 2,

"a therapist": 2,

"strattera": 2,

"medicine": 2,

"cbt": 2,

"ssris": 2,

"exercise": 2,

"hydroxyzine": 2,

"thepeloton": 1,

"group therapy": 1,

"self-behavioral treatment": 1,

"neuro feedback": 1,

"meditation": 1,

"cocaine": 1,

"mind games": 1,

"l-tyrosine": 1,

"the appropriate medication": 1,

"rehab": 1,

"ritilia n": 1,

"zoloft": 1,

"private assessment": 1,

"weight loss": 1,

"online therapy": 1,

"weed": 1,

"breast reduction surgery": 1,

"omega 3 supplements": 1,

"health care": 1,

"pain killer": 1,

"mental health care": 1,

"intima": 1,

"medications": 1,

"concert": 1,

"food": 1,

"resources": 1,

"psychiatr ist": 1,

"birth control pills": 1,

"counseling": 1,

"intervention": 1,

"cognitive behavioral therapy": 1,

"clonidine": 1,

"stimulant": 1,

"locybin": 1,

"nicotine": 1,

"art therapy": 1,

"online treatment": 1,

"stimulant medications": 1,

"mental health services": 1,

"magnesium": 1,

"amphetamine s": 1,

"natural remed": 1,

"mod af ini": 1,

"oral care": 1,

"alcohol": 1,

"digital therapy": 1,

"mental health": 1,

"menopause": 1,

"touch": 1,

"an effective therapist": 1,

"cigarettes": 1,

"the placebo": 1,

"rizatriptan": 1,

"a diagnosis": 1,

"various medications": 1,

"free tutoring program": 1,

"coffee": 1,

"pet s": 1,

"yoga": 1,

"internet": 1,

"anti-depressants": 1,

"sleep": 1,

"cannabis": 1,

"childhood treatment": 1,

"aerobic exercise": 1,

"the criteria": 1,

"an alternative treatment": 1,

"treatments": 1,

"a beta blocker": 1,

"an appointment": 1,

"advice": 1,

"in therapy": 1,

"guan fa cine": 1,

"breastfeeding": 1,

"emotional support": 1,

"a mood stabilize r": 1,

"hrt": 1,

"a stimulant": 1

}
