## Supplementary figures and images for "Linguistic Dynamics: Women vs. the General Population in Reddit’s ADHD Discussions"

### wordcloud_general.docx

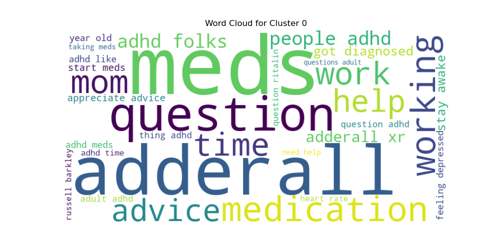

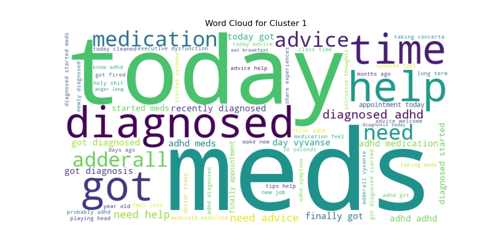

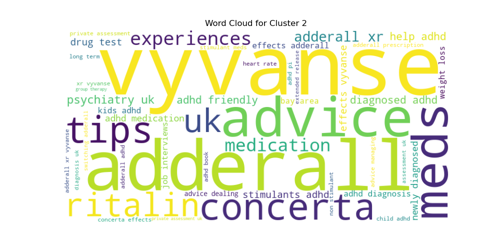

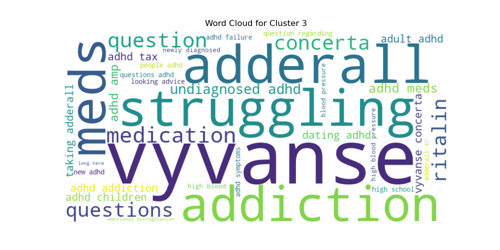

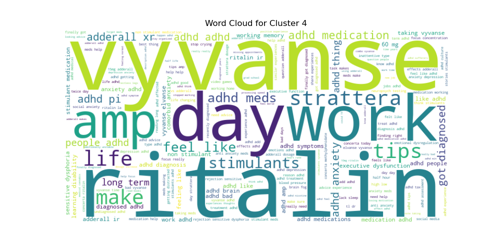

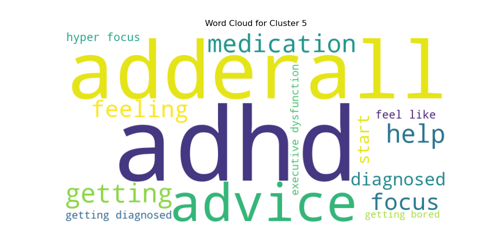

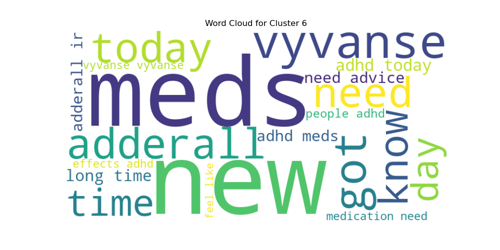

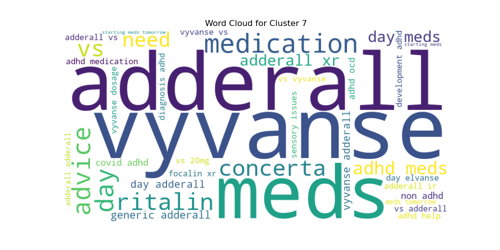

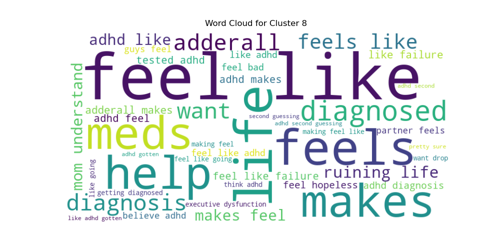

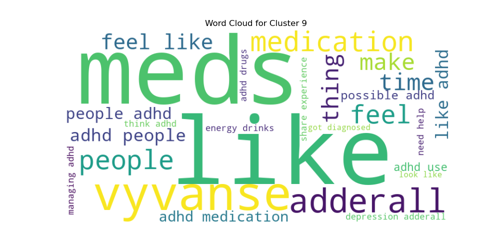

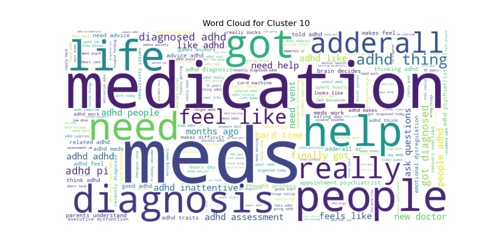

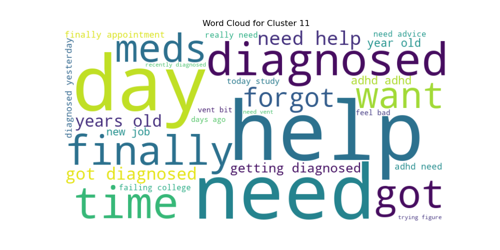

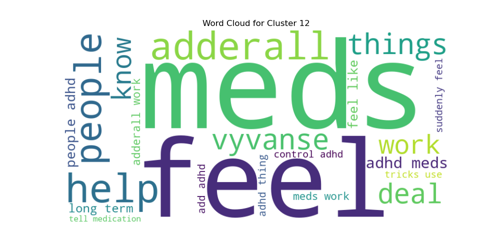

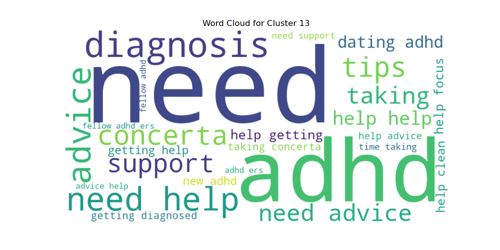

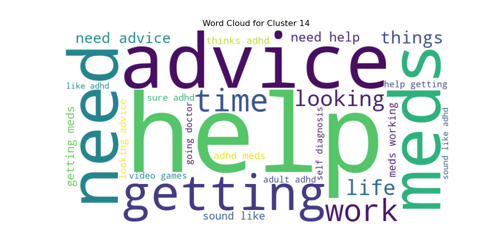

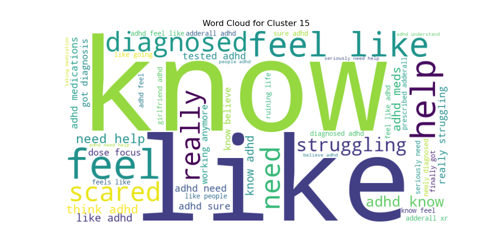

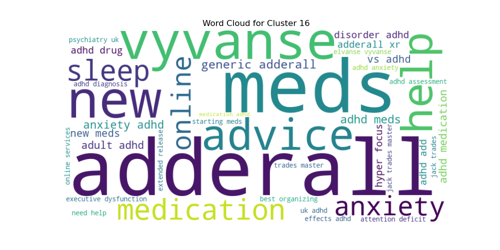

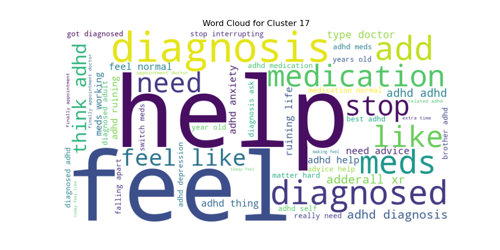

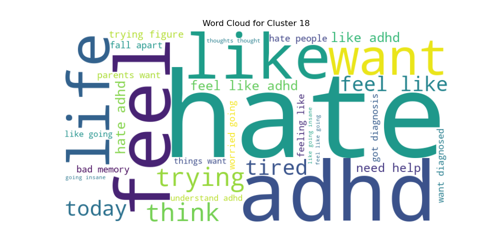

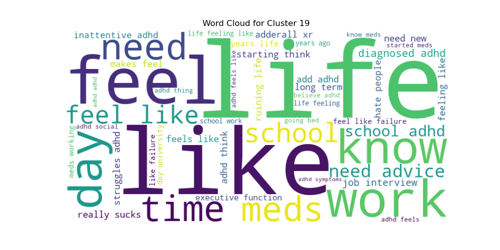

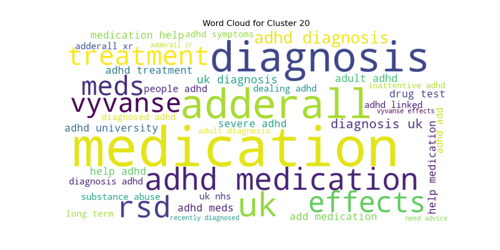

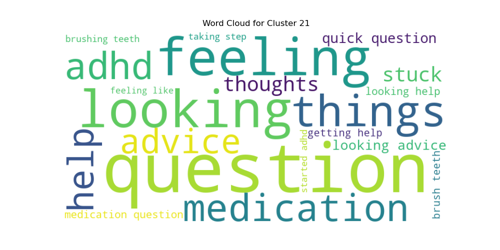

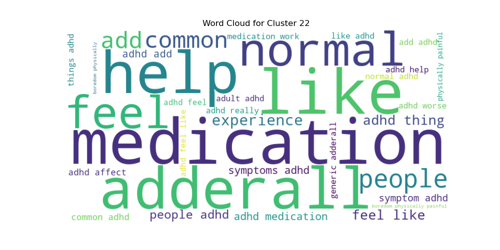

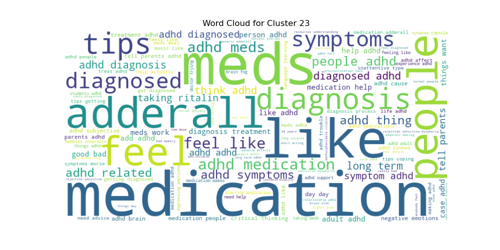

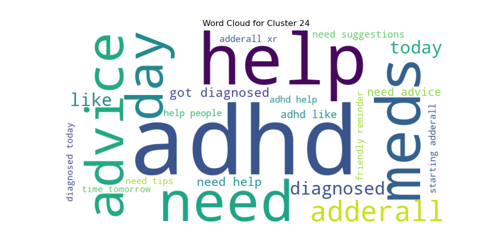

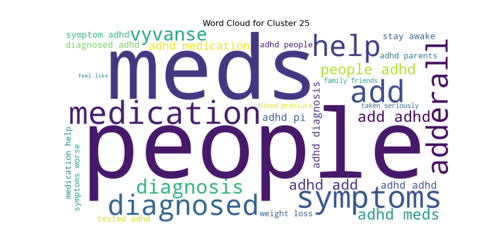

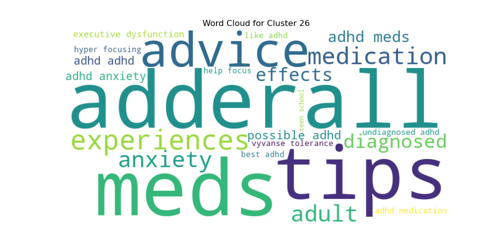

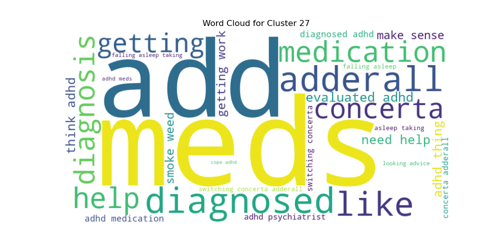

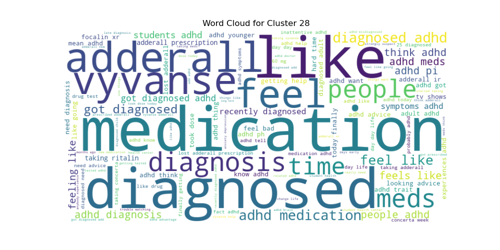

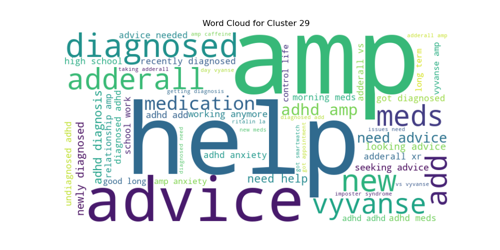

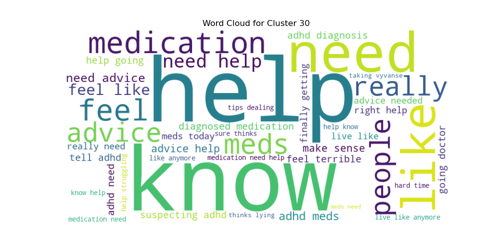

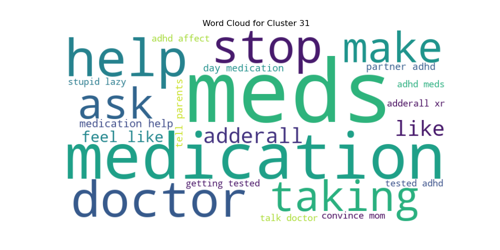

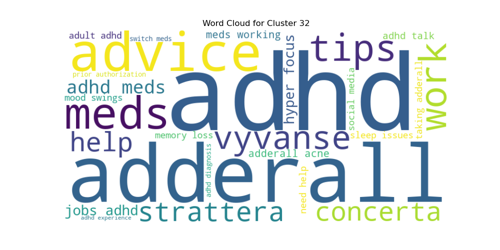

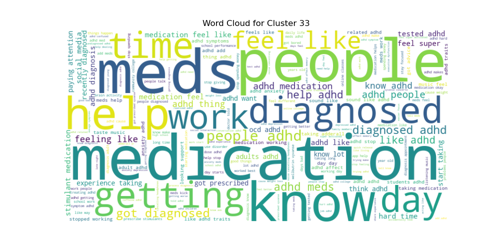

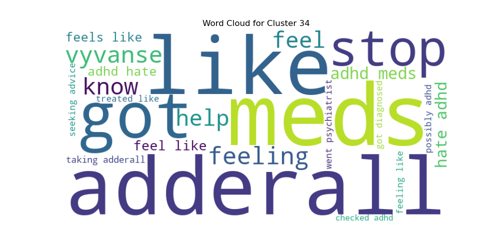

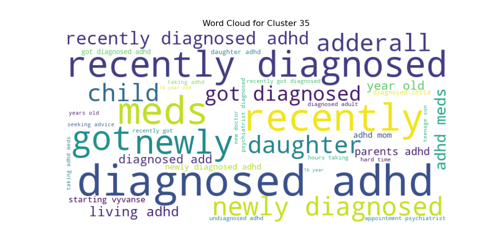

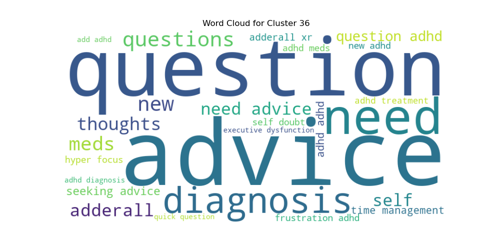

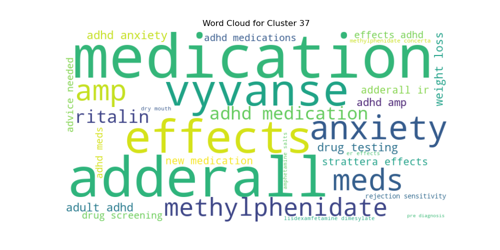

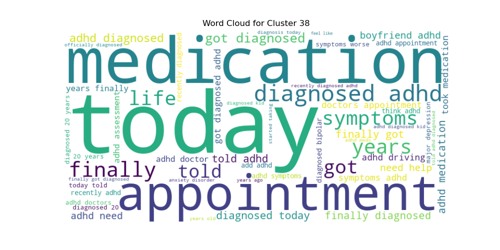

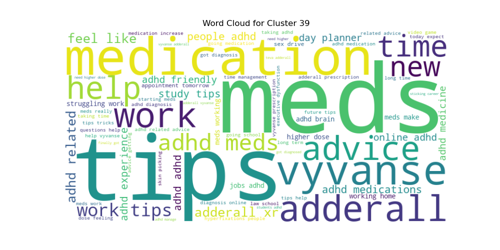

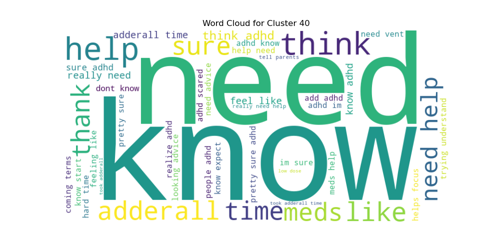

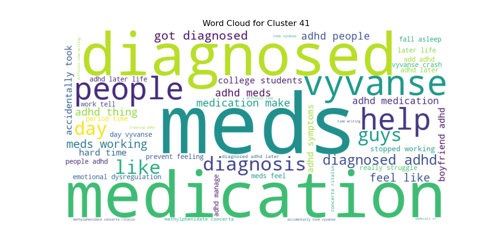

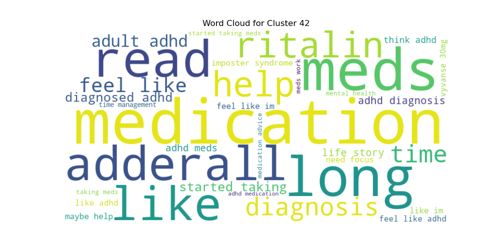

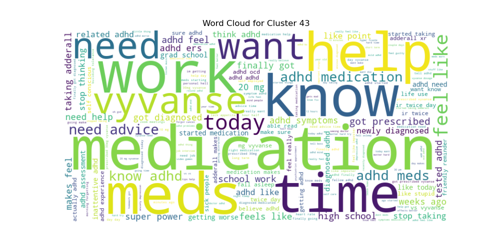

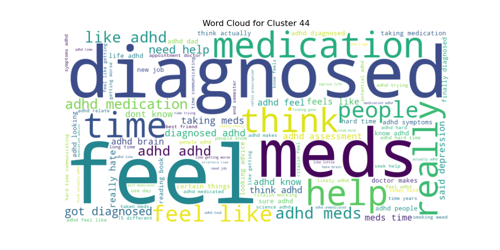

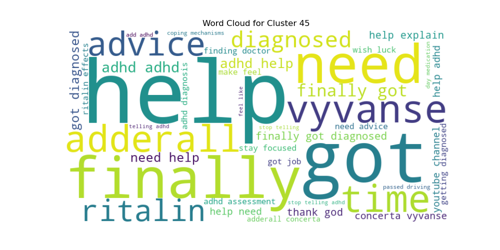

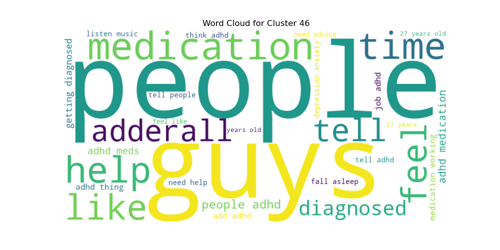

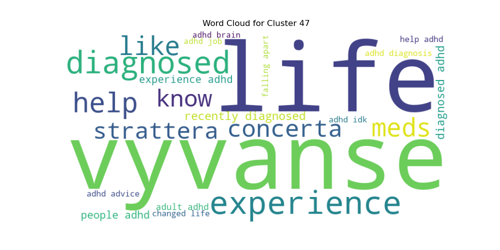

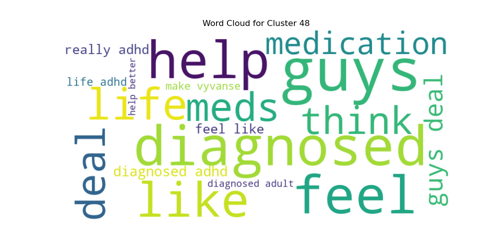

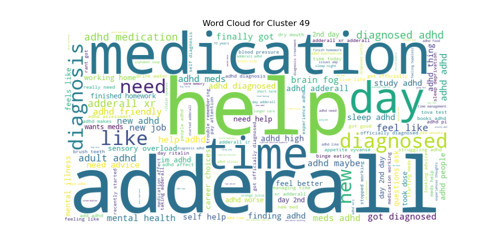

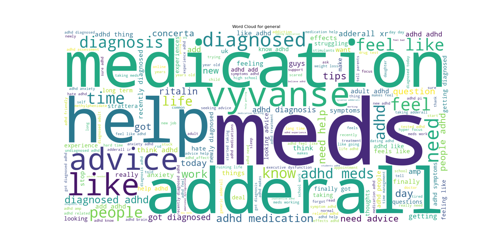

### wordcloud_women.docx

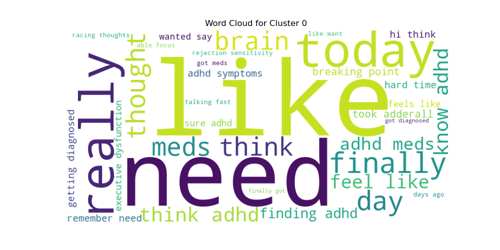

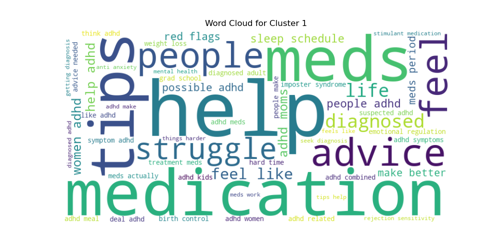

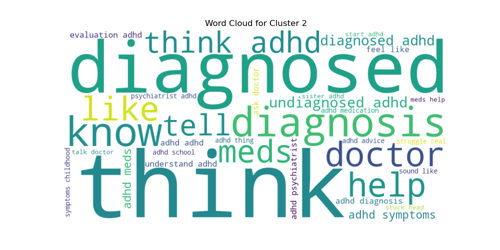

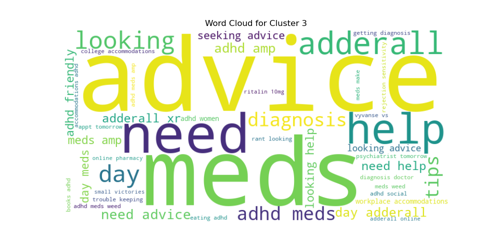

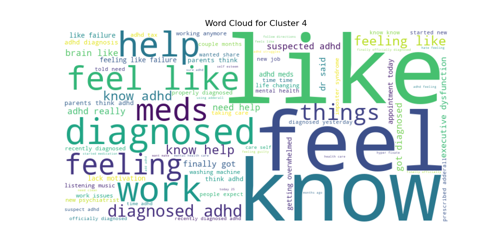

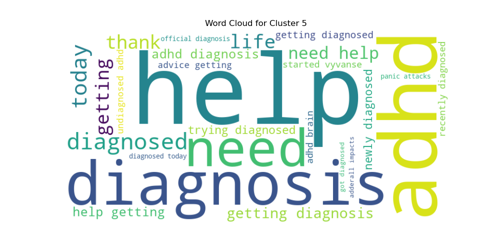

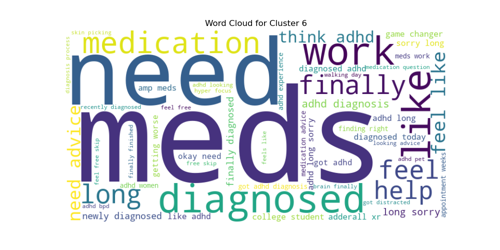

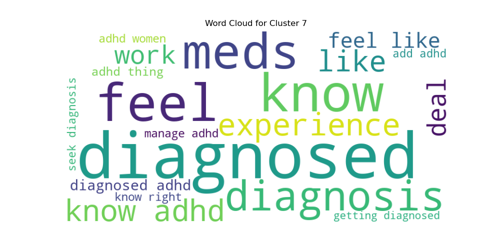

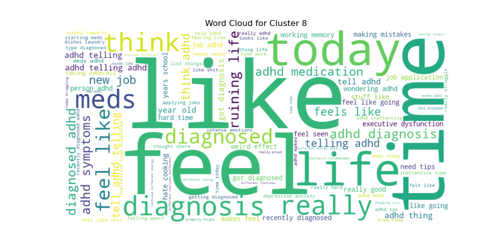

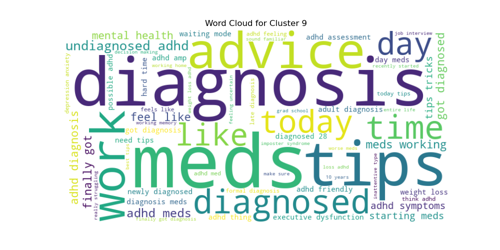

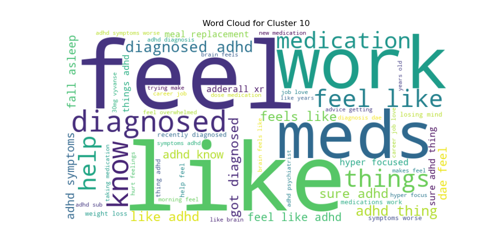

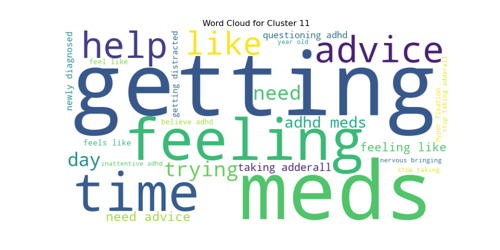

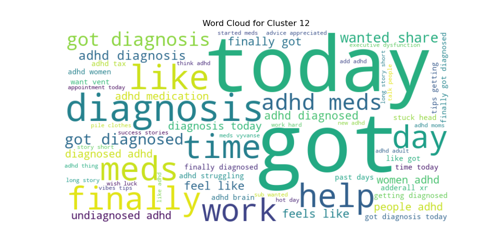

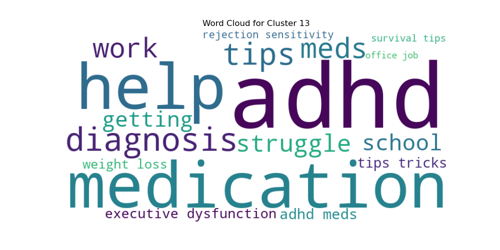

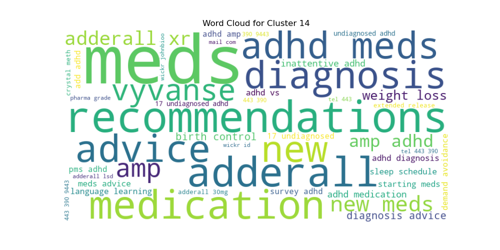

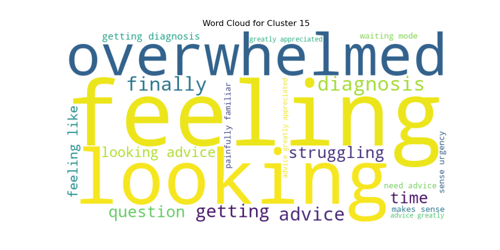

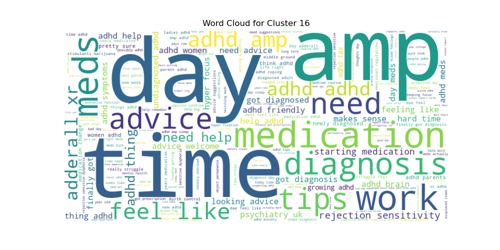

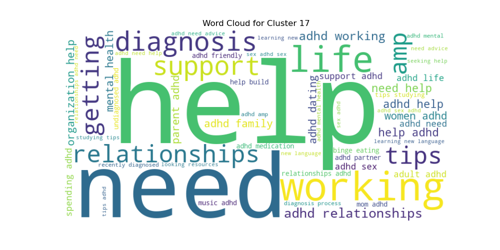

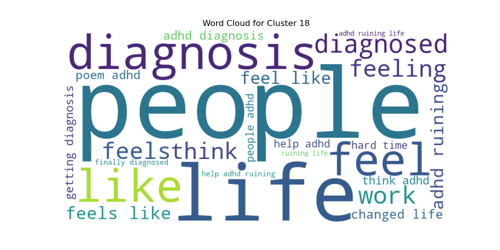

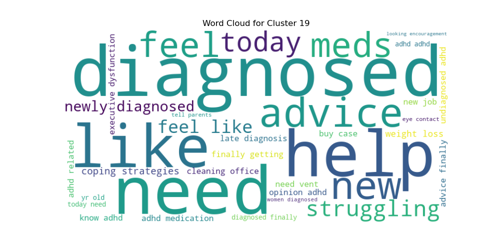

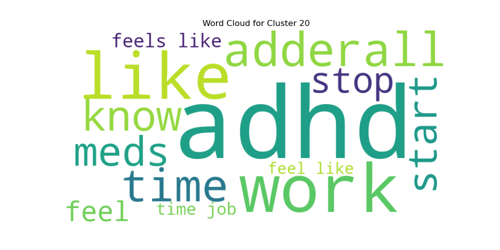

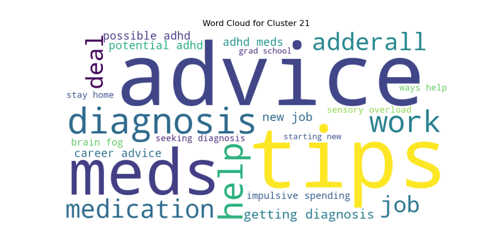

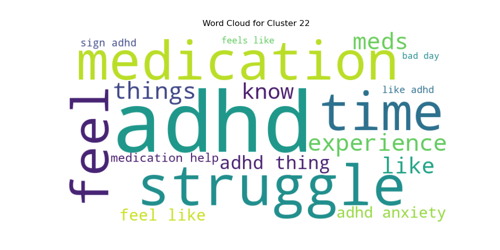

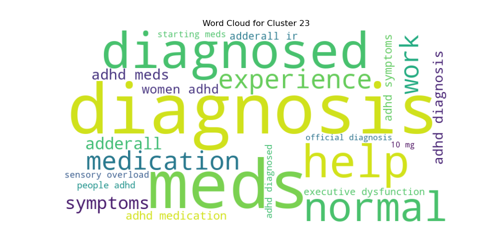

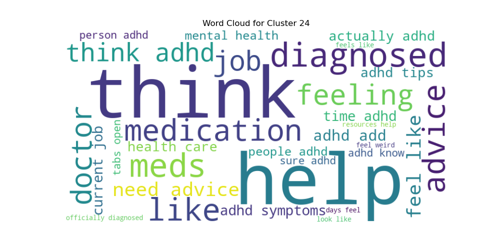

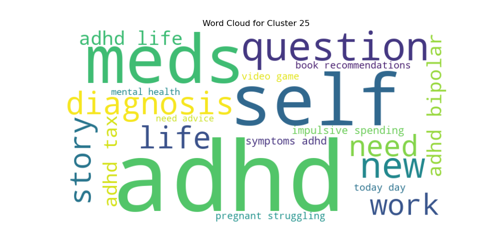

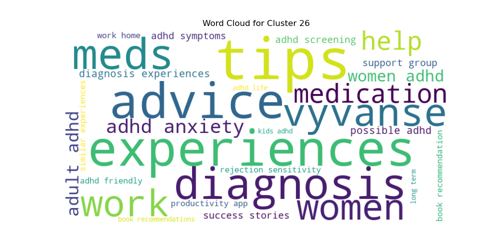

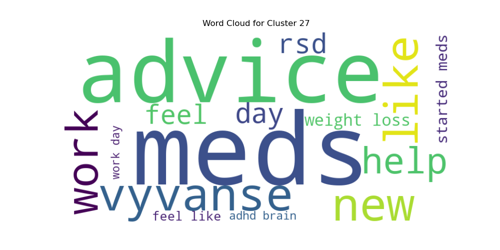

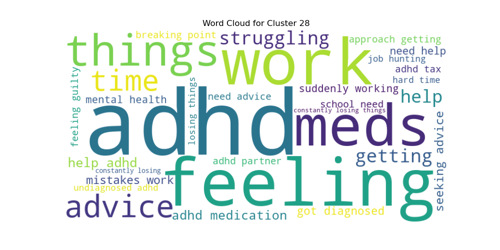

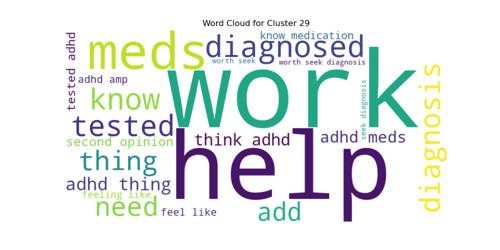

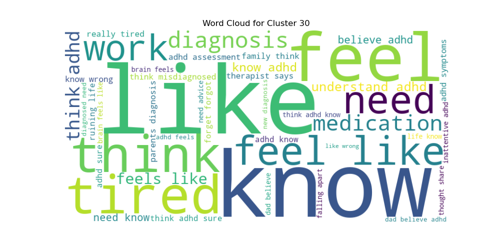

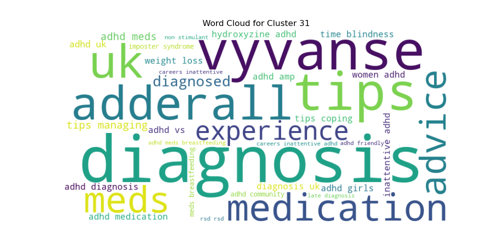

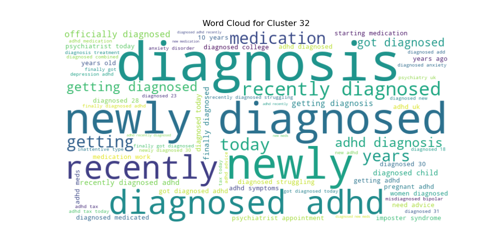

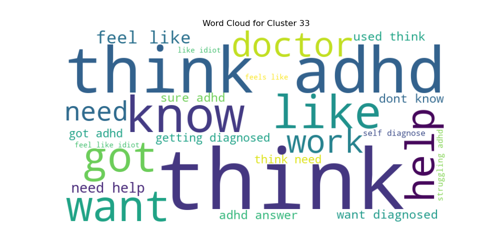

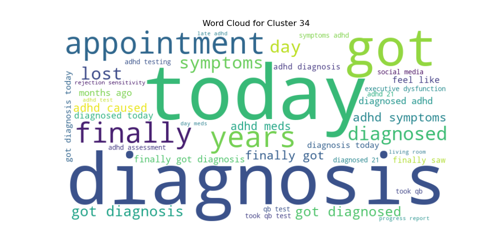

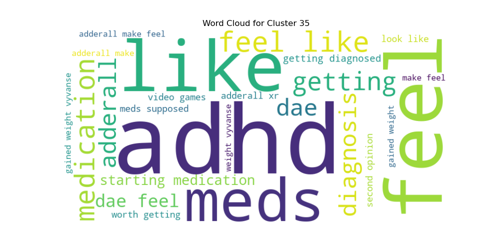

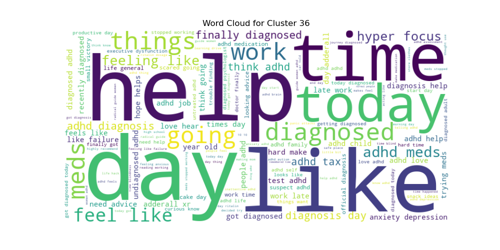

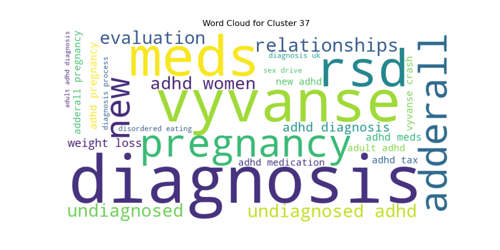

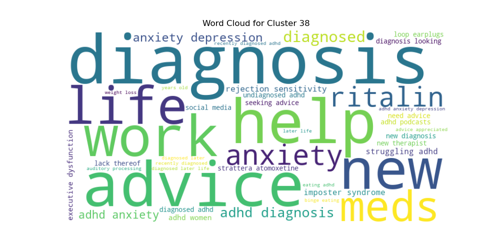

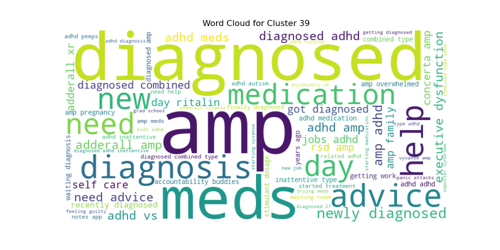

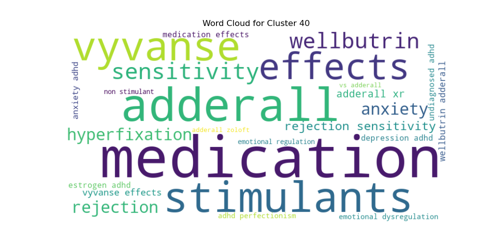

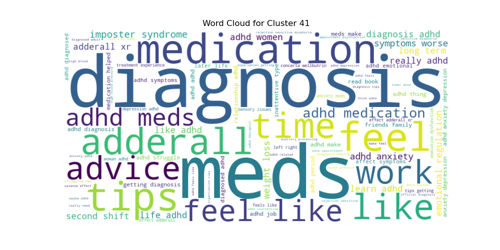

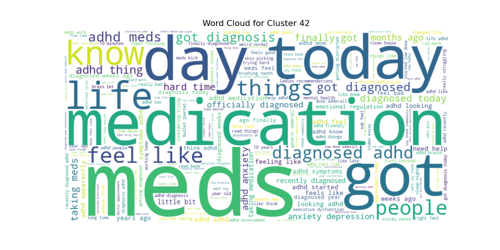

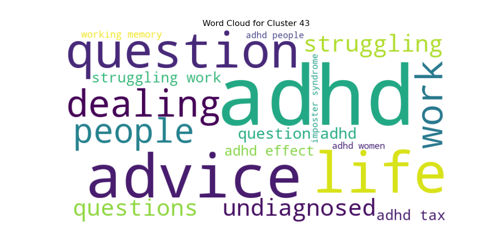

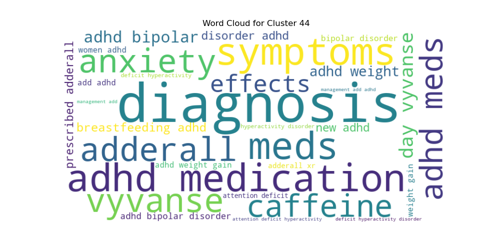

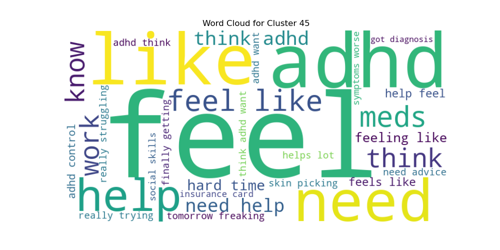

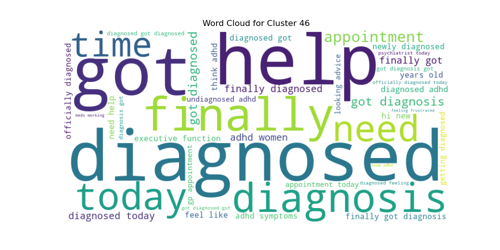

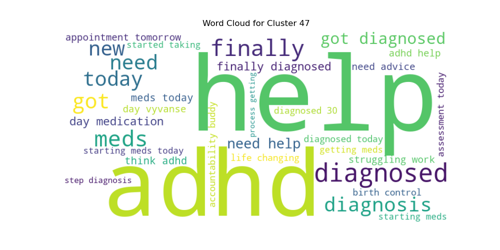

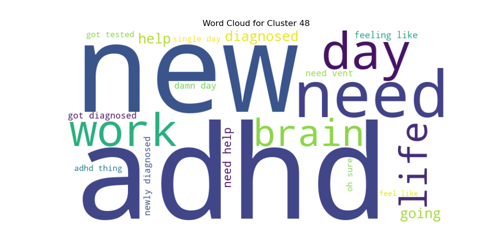

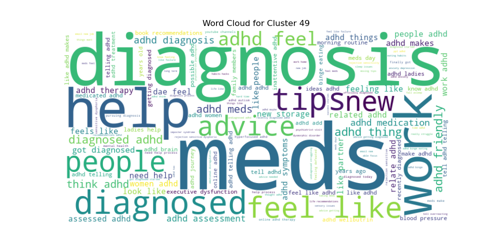

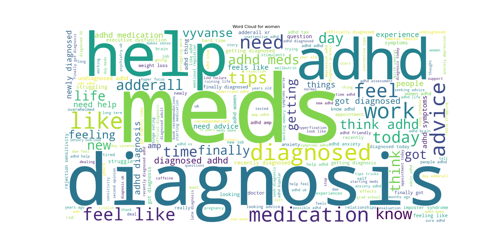
